# Untargeted metabolomics reveals sugar and homocysteine dysregulations in prodromal AD

**DOI:** 10.1101/2020.02.23.20025064

**Authors:** Ihab Hajjar, Chang Liu, Dean P. Jones, Karan Uppal

**Author notes:** Corresponding author: Ihab Hajjar, MD, MS, Associate Professor of Medicine and Neurology, Department of Neurology, Emory University, 6 Executive Park, Atlanta, GA 30329.

## Abstract

**Introduction:** Altered metabolism may occur early in Alzheimer’s disease (AD). We used untargeted high-resolution metabolomics in the cerebrospinal fluid (CSF) in mild cognitive impairment (MCI) to identify these alterations.

**Methods:** CSF from 92 normal controls and 93 MCI underwent untargeted metabolomics using high-resolution mass spectrometry with liquid chromatography. Partial least squares discriminant analysis was used followed by metabolite annotation and pathway enrichment analysis (PES). Significant features were correlated with disease phenotypes using spearman correlation.

**Results:** We identified 294 features differentially expressed between the 2 groups and 94 were annotated. PES showed that pathways related to sugar regulation (N-Glycan, *p*=0.0007; sialic acid, *p*=0.0014; Aminosugars, *p*=0.0042; galactose, *p*=0.0054) homocysteine regulation (*p*=0.0081) were differentially activated and significant features within these pathways correlated with disease phenotypes.

**Conclusion:** We identified a metabolic signature characterized by impairments in sugar and homocysteine regulation in prodromal AD. Targeting these changes may offer new therapeutic approaches to AD

**Research in Context:**

1. Systematic review: The authors searched PUBMED and Google Scholar for previous reports of metabolomics and Alzheimer’s disease. Search Terms included: mild cognitive impairment, Alzheimer’s disease “AND” metabolism, metabolomics. This search identified multiple small studies that have conducted untargeted metabolomics in AD. This search resulted in the following findings: Prior studies have either included small samples, used targeted approaches, or focused on plasma profiling. In this study, we conducted a case-control untargeted high resolution metabolomic study on the CSF of a larger sample of normal cognition and mild cognitive impairment.
2. Interpretation: We discovered that pathways in sugar metabolism, homocysteine and tyrosine were dysregulated in AD. Further, features that were significantly different between MCI and normal cognition had different patterns of association with cognitive, neuroimaging and Amyloid and tau biomarkers.
3. Future direction: These pathways offer new potential targets for AD

**Highlights:**

- Metabolic signature is detectable in prodromal AD
- Multiple sugar metabolism pathways are dysregulated in prodromal AD.
- S-adenosylmethionine is under- and S-adenosylhomocysteine is overexpressed in AD

## 1. Introduction

Alzheimer’s disease (AD) is characterized by a complex set of molecular pathways that begin decades before symptoms start.[1, 2] Changes in proteins, lipids and many other molecular networks have been described.[3, 4] The overlapping and interaction of these networks can obscure the root pathogenic mechanisms when not fully accounted for in molecular or analytical methods. The level of complexity in these networks is becoming more evident as the cumulative knowledge of AD pathogenesis has increased in the last decade. Disentangling these complexities is becoming more feasible due to the significant advances in high throughput technologies[5] coupled with novel bioinformatic tools including those developed by our team. [6] Recent examples applying the high throughput measurements of thousands of metabolites coupled with advanced bioinformatic approaches has comprehensively described molecular alterations and pathways in multiple diseases.[7-10] We apply these advanced in investigating underlying metabolic changes in AD.

Metabolomic research focuses on examining metabolites, small molecules (typically <1,500 Da), that are end products of multiple biological pathways and processes. The human metabolome is estimated to contain approximately 150,000 or more of such metabolites and a large fraction are still unidentified.[11] Metabolomics aids in identifying downstream perturbations from the genetic and post genetic pathways reflecting a functional signature of biochemical activities that are closer to the phenotypical changes.[12] Our work uses high resolution untargeted metabolomic approaches, which be a powerful tool in describing novel and previously unknown pathways involved in AD pathogenesis.

Brain hypometabolism has been reported symptomatic AD as well as before the onset of cognitive symptoms.[13] Preliminary studies have suggested the existence of multiple metabolic changes in this prodromal stage. [14, 15] However, many previous studies have either included small samples, used targeted approaches limited by prior knowledge, or focused on plasma profiling. In this study, we conducted a case-control untargeted high resolution metabolomic study on the CSF of a relatively larger sample of normal cognition (NC) and mild cognitive impairment (MCI), a prodromal state for AD. We aimed at investigating the alterations between NC and MCI in the metabolome and metabolic pathways using an established high resolution metabolomic biospecimen and data analysis pipeline. We further explored the association of these metabolic alterations with multiple disease phenotypes related to cognition, CSF Amyloid beta 1-42 (Aβ42) and tau biomarkers, and brain MRI measures.

## 2. Methods

### 2.1. Participants description

Data for the current analysis were drawn from the baseline assessment of participants in the Brain Stress Hypertension and Aging program (B-SHARP) at Emory University. B-SHARP participants undergo baseline cognitive assessments, neuroimaging, and lumbar punctures and are subsequently enrolled clinical studies. This analysis used data from the 185 participants enrolled from March 2016-January 2019 who had CSF obtained during their baseline evaluations. The protocol was approved by the Emory University Institutional Review Board prior to recruitment. Each participant provided a written informed consent.

The sample includes community-dwelling adults 50 years or older with NC or MCI. Potential study participants were identified either through a referral from the Goizueta Alzheimer’s Disease Research Center at Emory or through strategic community partnerships with grass root health education organizations, health fairs, advertisements and mail out announcements. An appropriate study informant, defined as an individual who has regular contact with the participant for at least once a week (in person or telephone), was also identified for each participant. The potential study participant attended a screening visit, during which they underwent cognitive testing. A study physician also performed a clinical evaluation, cognitive interview and a lumbar puncture.

### 2.2. Cognitive diagnosis and exclusionary criteria

Mild cognitive impairment (MCI) categorization was done using modified Peterson criteria. This modification included using the Montreal Cognitive Assessment (MoCA)[16] instead of Mini-Mental State Exam (MMSE).[17] MCI criteria included subjective memory complaints, a MoCA < 26, Clinical Dementia Rating (CDR) score, memory sum of boxes=0.5,[18] education adjusted cutoff score on Logical

Memory delayed recall of the Wechsler Memory Scale,[19] and preserved Functional Assessment Questionnaire (FAQ)<=7.[20] Normal cognition (NC) was defined as having no significant memory complaints beyond those expected for age, a MoCA score ≥26 points, a CDR score of 0 (including 0 on the Memory Box score), and preserved FAQ <=7. Participants were excluded if they had a history of stroke in the past three years, were unwilling or unable to undergo study procedures including MRI and LP, did not have a study informant, had a clinical diagnosis of dementia of any type, or abnormal serum Thyroid Stimulating Hormone (>10) or B12 (<250).

### 2.3. Cognitive Assessment and biomarker measurements

Demographics (age, sex, education), anthropometrics (weight and height), medical diagnosis, and medications were collected at baseline by interview. Cognitive assessment included those described above plus Trail Making Tests (Part A and B) a measure of executive function and Hopkins Verbal Learning test (HVLT) for episodic memory. Cognitive assessment was performed by trained personnel supervised by the study neuropsychologist. Following a fast of no less than 6 hours, CSF samples were collected via lumbar puncture using 24G Sprotte atraumatic spinal needles. Samples were collected in sterile polypropylene tubes, separated into 0.5cc aliquots and stored at −80 °C, Samples were subsequently shipped to and analyzed by the Biomarker Research Laboratory at the University of Pennsylvania (Dr. Leslie Shaw).[21] CSF Biomarkers: Aβ, t-Tau, and p-Tau were measured using the multiplex with the multiplex xMAP Luminex platform (Luminex Corp, Austin, TX) with Innogenetics (INNO-BIA AlzBio3; Ghent, Belgium; for research use–only reagents) immunoassay kit–based reagents. The test–retest reliabilities are 0.98, 0.90, and 0.85 for t-Tau, Aβ, and p-Tau181p, respectively. [21]

### 2.4. MRI Brain Imaging

Brain MRI’s were also completed at Emory University (3.0 Tesla Trio MRI scanner, Siemens Medical Solutions, Malvern, PA). Anatomical images were acquired using high-resolution three-dimensional (3D) magnetization-prepared rapid acquisition with gradient echo (MPRAGE). Images were then digitally saved for offline processing. Hippocampal volume and other volumetric measurements were calculated using the free-surfer package with manual supervision. Quality checks were performed for each scan. Left and right hippocampal volumes were obtained and combined to derive the total hippocampal volume and cortical thickness. Intra-cranial volume (ICV, mm^3) was also derived from this analysis. Volumetric measurements using free surfer has been shown to provide similar estimates to a fully manual procedure.[22] We used ICV-adjusted hippocampal volume to reflect the degree of neurodegeneration for each participant.[23]

### 2.5. Untargeted metabolomic High-resolution metabolomics (HRM) approaches and pipeline

Our metabolomic approaches used an established pipeline developed at the Clinical Biomarker Laboratory, led by Dr. Dean Jones (diagrammatic representation of this pipeline is included in the online supplement). HRM was completed using established methods by an analyst blinded to sample identity.[8, 24] Briefly, CSF samples were prepared and analyzed in batches of 20. Prior to analysis, CSF aliquots were removed from storage at −80°C and thawed on ice. A 65 μL aliquot of CSF was then treated with 130 μL of LC-MS grade acetonitrile, equilibrated for 30 min on ice and centrifuged (16.1 ×g at 4°C) for 10 minutes to remove precipitated proteins. The supernatant was added to an autosampler vial and maintained at 4°C until analysis. Sample extracts were analyzed using liquid chromatography (LC) and Fourier transform high-resolution mass spectrometry (Dionex Ultimate 3000, Q-Exactive HF, Thermo Scientific). For each sample, 10 μL aliquots were analyzed in triplicate using hydrophilic interaction liquid chromatography (HILIC) with electrospray ionization (ESI) source operated in positive mode. This use of complementary chromatography phases and ionization polarity has been shown to improve detection of endogenous and exogenous chemicals.[25] Analyte separation was accomplished by HILIC using a 2.1 mm × 100 mm × 2.6 μm Accucore HILIC column (Thermo Scientific) and an eluent gradient (A= 2% formic acid, B= water, C= acetonitrile) consisting of an initial 1.5 min period of 10% A, 10% B, 80% C, followed by linear increase to 10% A, 80% B, 10% C at 6 min and then held for an additional 4 min, resulting in a total runtime of 10 min per injection. Mobile phase flow rate was held at 0.35 mL/min for the first 1.5 min, increased to 0.5 mL/min and held for the final 4 min.

The high-resolution mass spectrometer was operated in full scan mode at 120,000 resolution and mass-to-charge ratio (*m/z*) range 85–1275. Probe temperature, capillary temperature, sweep gas and S-Lens RF levels were maintained at 200°C, 300°C, 1 arbitrary units (AU), and 45 AU, respectively, for both polarities. Positive tune settings for sheath gas, auxiliary gas, sweep gas and spray voltage setting were 45 AU, 25 AU and 3.5 kV, respectively. Raw data files were extracted and aligned using apLCMS[26] with modifications by xMSanalyzer.[27] Uniquely detected ions consisted of accurate mass *m/z*, retention time and ion abundance, referred to as *m/z* features. Data filtering was performed to remove *m/z* features with median coefficient of variation (CV) within technical replicates ≥ 75%. Additionally, only samples with Pearson correlation within technical replicates ≥ 0.7 were used for downstream analysis. Feature intensities for triplicates were median summarized with the requirement that at least two replicates had non-missing values. Batch-effect correction was performed using ComBat[28].

### 2.6. Metabolome-wide association analysis (MWAS)

A feature was retained for further analysis if at least 90% of the subjects had non-zero intensity reading in either MCI or NC groups. After exclusion, the missing values for a feature were imputed as half of the lowest signal detected for that feature across all samples. Following data filtering, all intensity values were log_2_ transformed to reduce heteroscedasticity and quantile normalized to reduce systematic errors due to technical and other non-biological factors. MWAS was conducted using partial least squares discriminant analysis (PLS-DA) implemented in the mixOmics[29] R package and features were selected based on the variable importance for projection (VIP) criteria. P-values were obtained for each feature using a permutation test. A 1000-permutation approach was performed by randomly shuffling the group labels of subjects and performing feature selection using PLS-DA at each iteration.^30^ Multiple testing correction was performed using Storey and Tibshirani FDR adjustment.[30] Discriminatory features were selected using the thresholds of variable importance for projection (VIP) ≥ 2, permutation derived *p*<0.05, and FDR< 0.1. Only features that passed all three criteria were considered significantly different between the two groups. Manhattan plot was used to visualize the pattern of differential expression across all features with respect to molecular mass. Fold change of log_2_ transformed intensity values was calculated for each feature as the difference between the average intensity of the two groups, log_2_ FC=average_NC_-average_MCI_.

### 2.7. Pathway analysis

Pathway enrichment analysis was performed using mummichog (v2.0.6), which uses both *m/z* and retention time, and included discriminatory features that met the following criteria: VIP ≥1.5, *p*<0.05, and FDR<0.1. A lower VIP was used to increase enrichment within the pathway and prevent information loss.[10, 31] Detailed descriptions of mummichog computational procedures were previously published for V1.0.[32] Discriminatory features detected in the pathways were further tested for differential expression between the NC and MCI groups using Wilcoxon Rank Sum test.

### 2.8. Metabolite annotation and identification

Metabolite annotation and identification was performed using MS/MS, comparison with in-house library of confirmed metabolites, and using xMSannotator^33^ with the Human Metabolome Database^**34**^ (HMDB). Discriminatory features that were associated with the significantly enriched pathways and had *p*<0.05 using the Wilcoxon Rank Sum test were selected for MS/MS analysis. For MS/MS, samples were analyzed using a Thermo Fusion Orbitrap high-resolution (120,000 mass resolution) mass spectrometer (Thermo Fisher Scientific, San Diego, CA) operated in positive ion mode with 5-minute HILIC column chromatography and similar source conditions used for the untargeted metabolic profiling. Prior to analysis, CSF proteins were precipitated using acetonitrile:water (2:1 vol/vol) and allowed to sit on ice for 30 minutes. The supernatant was then carefully pipetted for MS/MS analysis. The tandem mass spectrometry data was processed using the *xcmsSet* and *xcmsFragments* functions in XCMS[33] to extract the MS/MS fragments associated with each parent mass and the experimental spectra were compared with in-silico fragmentation using MetFrag[34] or the spectra available from mzCloud (https://www.mzcloud.org/).

We further annotated and confirmed identities of the selected metabolites using an in-house library of metabolites that have been previously confirmed by comparing the retention time and MS/MS of the metabolic feature with authentic standards. Additionally, we performed computational annotation using xMSannotator^**33**^ (v1.3.2) with the HMDB^**34**^ (v3.5). xMSannotator uses adduct/isotope patterns, correlation in intensities across all samples, retention time difference between adducts/isotopes of a metabolite, and network and pathway associations for associating *m/z* features with known metabolites and categorizing database matches into different confidence levels.[35] This multi-step annotation process reduces the number of false matches as compared to only *m/z*-based database search. Metabolite identification levels were assigned using an adapted version of the criteria proposed by Schymanski et al.: a) confirmed by MS/MS and co-elution with authentic standards (level 1); b) confirmed by MS/MS and matches with online databases or in-silico predicted spectra (level 2); c) confirmed by MS/MS at the chemical class level, but no evidence for a specific metabolite (level 3); d) computationally assigned annotation using xMSannotator (medium or high confidence) (level 4); e) accurate mass match (level 5).[36]

### 2.9. Association of discriminatory features with other disease phenotypes

Discriminatory metabolites associated with significantly enriched pathways were then tested for associations with three AD phenotypical or endophenotypic areas: cognitive performance (MoCA for global function, TMT A and B for executive function and HVLT-delayed recall for episodic memory), neuroimaging (hippocampal volume and cortical thickness as indicators of neurodegeneration) and CSF AD biomarkers (Amyloid beta 1-42 (Aβ42), total and phosphorylated tau (tau, Ptau) using Spearman’s correlation analyses. A heatmap was used to visualize the correlation patterns between significant metabolic features and these measures.

## 3. Results

### 3.1 Participant

Of the 185 participants who provided CSF, 93 were MCI and 92 were normal controls. The basic clinical characteristics of the sample are provided in **Table 1**. The MCI group were older (*p*=0.007) and had higher levels of Tau and p-tau (both *p*<0.0001), but not Abeta (*p*=0.6). They also had lower cognitive performance in all measures as expected and lower hippocampal volume (*p*<0.0001).

**Table 1.**
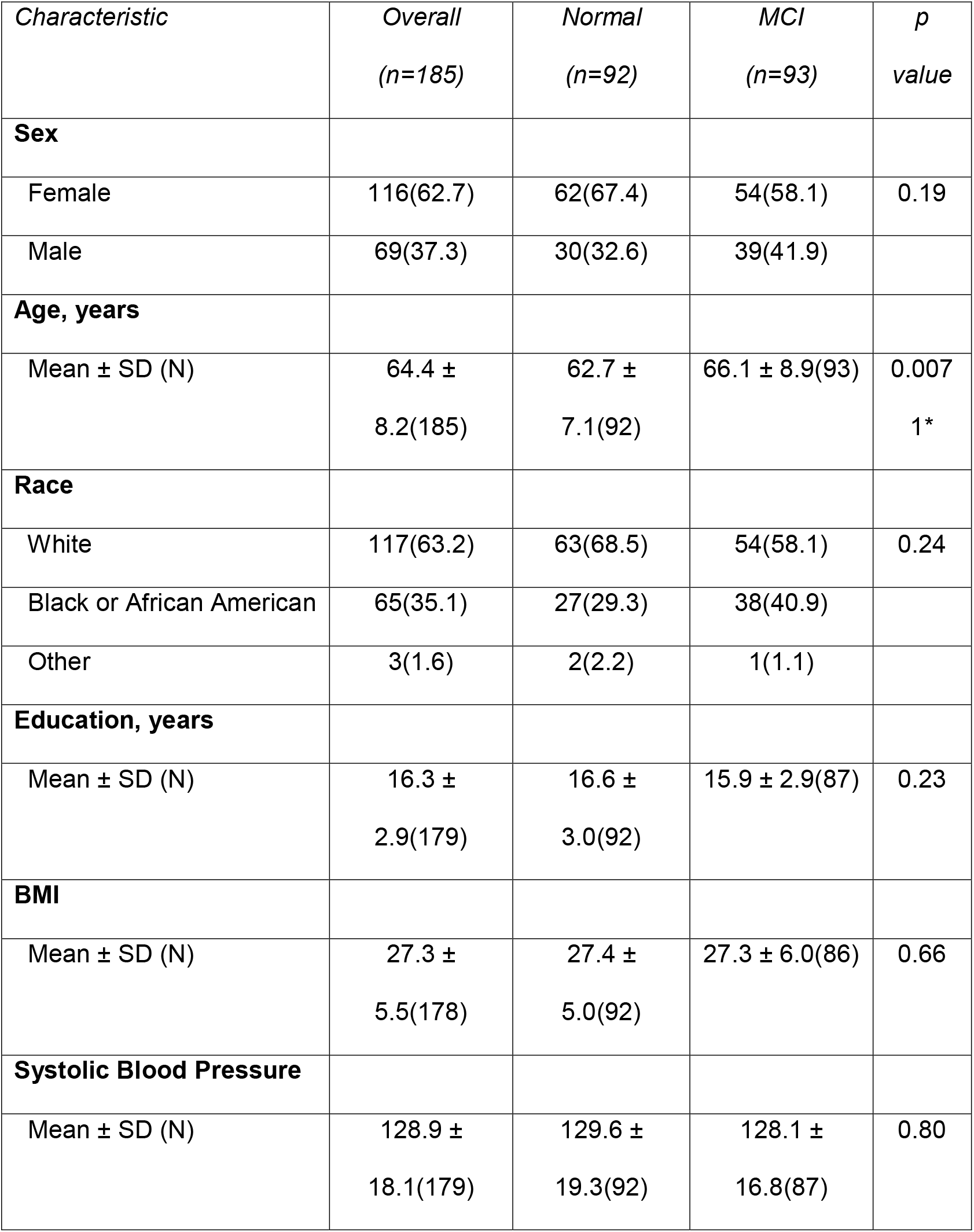
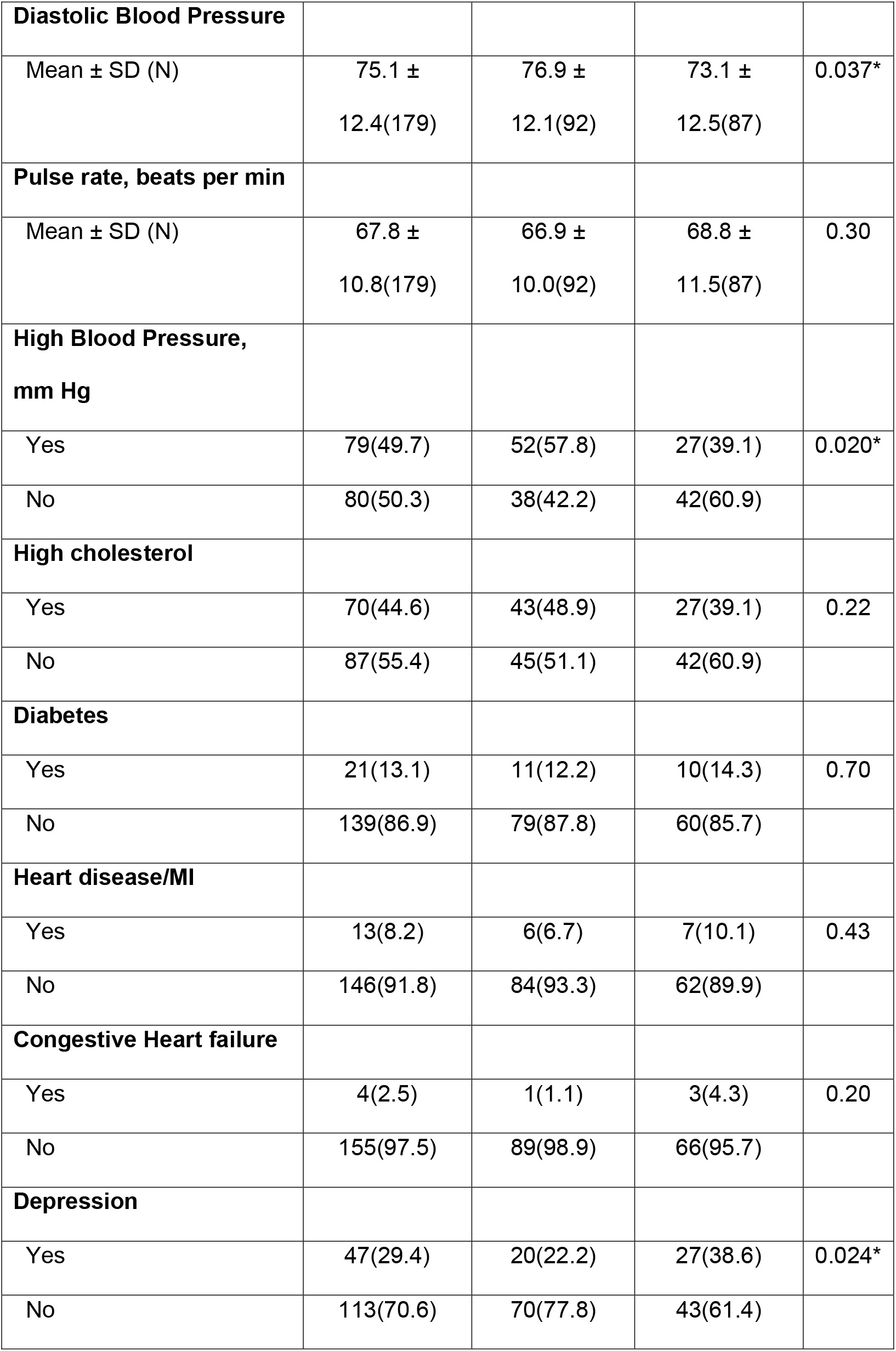
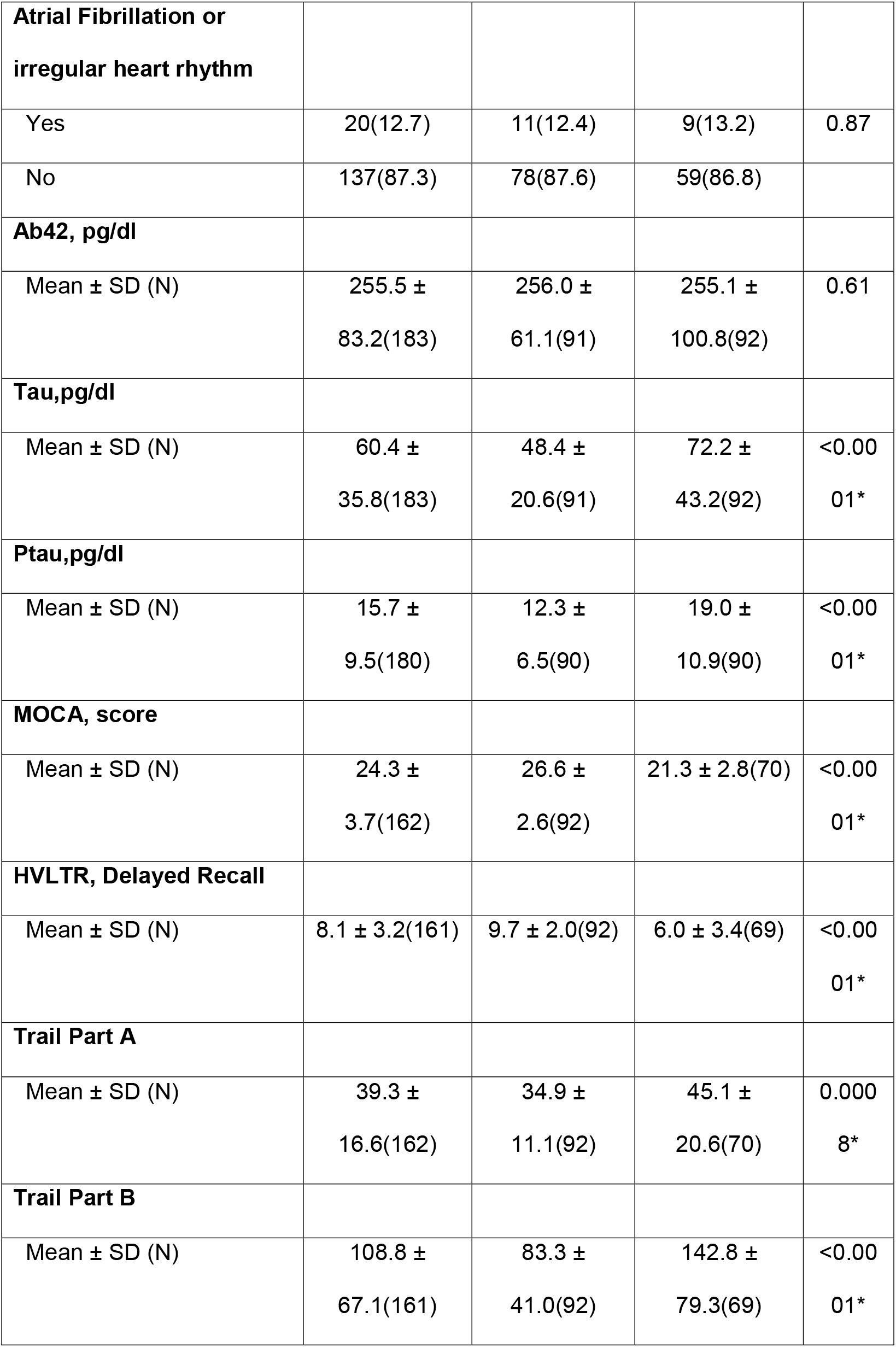
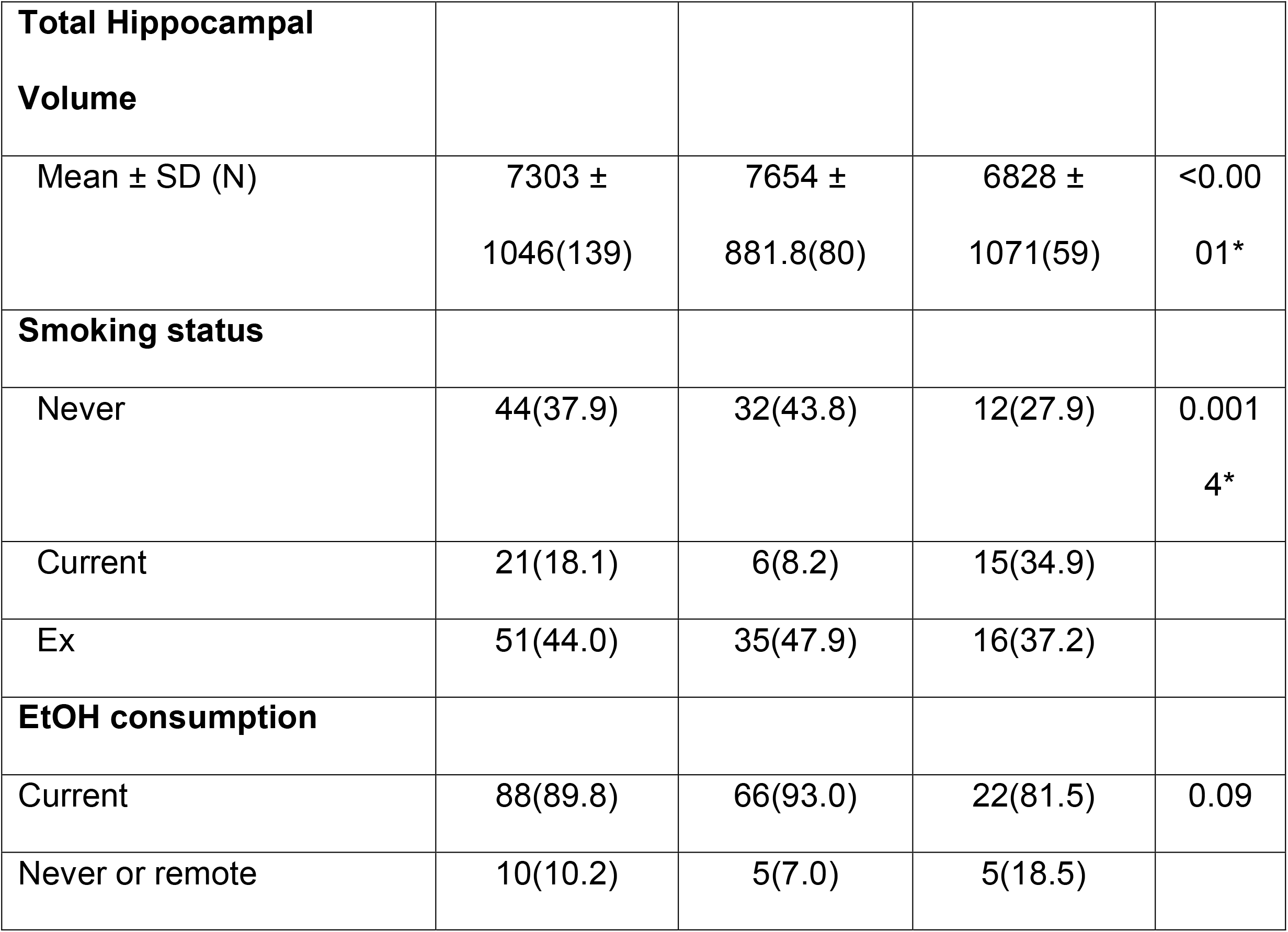
Characteristics of the overall sample by the 2 groups, Normal Cognition and Mild Cognitive Impairment (MCI)

| *Characteristic* | *Overall*<br>*(n=185)* | *Normal*<br>*(n=92)* | *MCI*<br>*(n=93)* | *p*<br>*value* |
| --- | --- | --- | --- | --- |
| **Sex** |  |  |  |  |
| Female | 116(62.7) | 62(67.4) | 54(58.1) | 0.19 |
| Male | 69(37.3) | 30(32.6) | 39(41.9) |  |
| **Age, years** |  |  |  |  |
| Mean ± SD (N) | 64.4 ±<br>8.2(185) | 62.7 ±<br>7.1(92) | 66.1 ± 8.9(93) | 0.007<br>1* |
| **Race** |  |  |  |  |
| White | 117(63.2) | 63(68.5) | 54(58.1) | 0.24 |
| Black or African American | 65(35.1) | 27(29.3) | 38(40.9) |  |
| Other | 3(1.6) | 2(2.2) | 1(1.1) |  |
| **Education, years** |  |  |  |  |
| Mean ± SD (N) | 16.3 ±<br>2.9(179) | 16.6 ±<br>3.0(92) | 15.9 ± 2.9(87) | 0.23 |
| **BMI** |  |  |  |  |
| Mean ± SD (N) | 27.3 ±<br>5.5(178) | 27.4 ±<br>5.0(92) | 27.3 ± 6.0(86) | 0.66 |
| **Systolic Blood Pressure** |  |  |  |  |
| Mean ± SD (N) | 128.9 ±<br>18.1(179) | 129.6 ±<br>19.3(92) | 128.1 ±<br>16.8(87) | 0.80 |
| **Diastolic Blood Pressure** |  |  |  |  |
| Mean ± SD (N) | 75.1 ±<br>12.4(179) | 76.9 ±<br>12.1(92) | 73.1 ±<br>12.5(87) | 0.037* |
| **Pulse rate, beats per min** |  |  |  |  |
| Mean ± SD (N) | 67.8 ±<br>10.8(179) | 66.9 ±<br>10.0(92) | 68.8 ±<br>11.5(87) | 0.30 |
| **High Blood Pressure,**<br>**mm Hg** |  |  |  |  |
| Yes | 79(49.7) | 52(57.8) | 27(39.1) | 0.020* |
| No | 80(50.3) | 38(42.2) | 42(60.9) |  |
| **High cholesterol** |  |  |  |  |
| Yes | 70(44.6) | 43(48.9) | 27(39.1) | 0.22 |
| No | 87(55.4) | 45(51.1) | 42(60.9) |  |
| **Diabetes** |  |  |  |  |
| Yes | 21(13.1) | 11(12.2) | 10(14.3) | 0.70 |
| No | 139(86.9) | 79(87.8) | 60(85.7) |  |
| **Heart disease/MI** |  |  |  |  |
| Yes | 13(8.2) | 6(6.7) | 7(10.1) | 0.43 |
| No | 146(91.8) | 84(93.3) | 62(89.9) |  |
| **Congestive Heart failure** |  |  |  |  |
| Yes | 4(2.5) | 1(1.1) | 3(4.3) | 0.20 |
| No | 155(97.5) | 89(98.9) | 66(95.7) |  |
| **Depression** |  |  |  |  |
| Yes | 47(29.4) | 20(22.2) | 27(38.6) | 0.024* |
| No | 113(70.6) | 70(77.8) | 43(61.4) |  |
| **Atrial Fibrillation or irregular heart rhythm** |  |  |  |  |
| Yes | 20(12.7) | 11(12.4) | 9(13.2) | 0.87 |
| No | 137(87.3) | 78(87.6) | 59(86.8) |  |
| **Ab42, pg/dl** |  |  |  |  |
| Mean ± SD (N) | 255.5 ±<br>83.2(183) | 256.0 ±<br>61.1(91) | 255.1 ±<br>100.8(92) | 0.61 |
| **Tau,pg/dl** |  |  |  |  |
| Mean ± SD (N) | 60.4 ±<br>35.8(183) | 48.4 ±<br>20.6(91) | 72.2 ±<br>43.2(92) | <0.00<br>01* |
| **Ptau,pg/dl** |  |  |  |  |
| Mean ± SD (N) | 15.7 ±<br>9.5(180) | 12.3 ±<br>6.5(90) | 19.0 ±<br>10.9(90) | <0.00<br>01* |
| **MOCA, score** |  |  |  |  |
| Mean ± SD (N) | 24.3 ±<br>3.7(162) | 26.6 ±<br>2.6(92) | 21.3 ± 2.8(70) | <0.00<br>01* |
| **HVLTR, Delayed Recall** |  |  |  |  |
| Mean ± SD (N) | 8.1 ± 3.2(161) | 9.7 ± 2.0(92) | 6.0 ± 3.4(69) | <0.00<br>01* |
| **Trail Part A** |  |  |  |  |
| Mean ± SD (N) | 39.3 ±<br>16.6(162) | 34.9 ±<br>11.1(92) | 45.1 ±<br>20.6(70) | 0.000<br>8* |
| **Trail Part B** |  |  |  |  |
| Mean ± SD (N) | 108.8 ±<br>67.1(161) | 83.3 ±<br>41.0(92) | 142.8 ±<br>79.3(69) | <0.00<br>01* |
| **Total Hippocampal Volume** |  |  |  |  |
| Mean ± SD (N) | 7303 ±<br>1046(139) | 7654 ±<br>881.8(80) | 6828 ±<br>1071(59) | <0.00<br>01* |
| **Smoking status** |  |  |  |  |
| Never | 44(37.9) | 32(43.8) | 12(27.9) | 0.001<br>4* |
| Current | 21(18.1) | 6(8.2) | 15(34.9) |  |
| Ex | 51(44.0) | 35(47.9) | 16(37.2) |  |
| **EtOH consumption** |  |  |  |  |
| Current | 88(89.8) | 66(93.0) | 22(81.5) | 0.09 |
| Never or remote | 10(10.2) | 5(7.0) | 5(18.5) |  |

### 3.2 MWAS results

Overall, 13,064 features were detected, and 8,043 features met the data filtering criteria and were used for downstream analyses. Using partial least squares discriminant analysis (PLS-DA), 294 discriminatory features were identified using the predefined criteria (**Figure 1**). Of those, 107 features were under-expressed and 187 features were overexpressed in MCI patients relative to NC, as shown in Figure 1. Of the 294 features, 94 were successfully matched to known metabolites in HMDB using xMSannotator with an annotation confidence score of medium or high (**Supplemental Table 1**).

**Figure 1:**
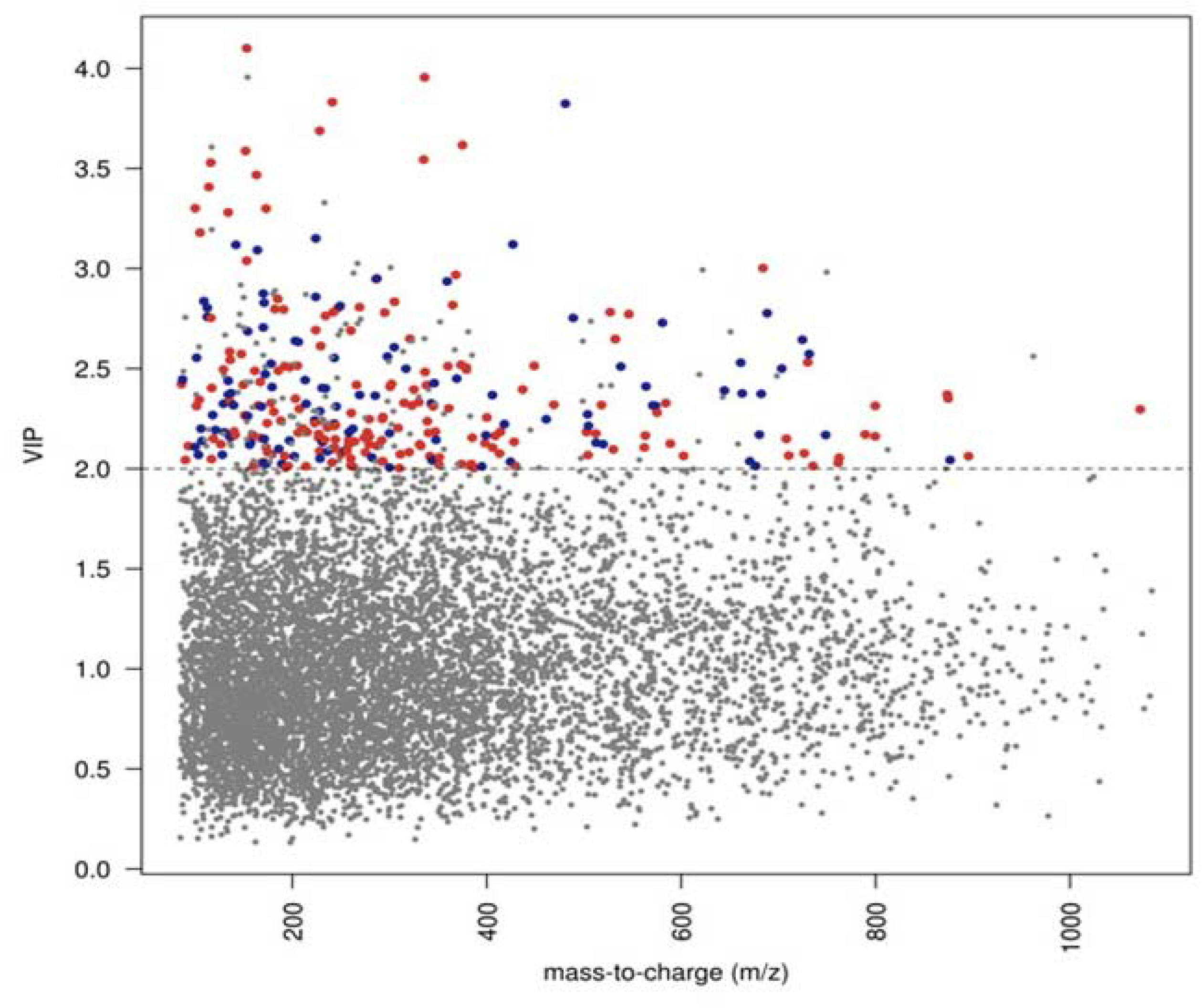
Manhattan plot shows the VIP and *m/z* of 8,043 features. A total of 294 features were significantly different between MCI cases (n=93) and controls (n=92) by PLS-DA using a VIP measure of 2.0 or greater (threshold indicated by horizontal line). 187 metabolic features increased (red dots) and 107 decreased (blue dots) in MCI patients compared to controls are indicated.

### 3.3 Pathway Analysis

To enhance the coverage of metabolites for pathway enrichment analyses and to prevent information loss, 1,049 discriminatory features were included using the less stringent criteria of VIP>1.5, *p*<0.05, and FDR<0.1. We identified 13 pathways that were perturbed between the MCI and normal control groups which are shown in **Figure 2**. The top 4 pathways were related to bioenergetics and glucose metabolism: N-Glycan (*p*=0.0007), Sialic Acid (*p*=0.0014), Amino-sugars (*p*=0.0042), and Galactose (*p*=0.0054) metabolism. Keratan sulfate (*p*=0.0173), Methionine (*p*=0.0081), Cyanocobalamin (*p*=0.0106), Tyrosine (*p*=0.0193), Purine (*p*=0.0352) and Biopterine (*p*=0.0275) were also differentially activated between the 2 groups. Within the enriched pathways that were significantly different between NC and MCI, multiple features with an identification confidence of 1 to 5 were differentially expressed and are shown in **Table 2**. Combined together, the overall CSF metabolic signature for MCI is shown in the KEGG pathway map 01100 (metabolic pathways; Homo sapiens) and the corresponding boxplots are shown in **Figure 3**. This signature is characterized by increased expressions of features related to sugar metabolism/bioenergetics, homocysteine, Tyrosine and Biopterin pathways and lower expression of methionine. The complete list of features in these analyses is provided in **Supplementary Table 2**.

**Table 2:**
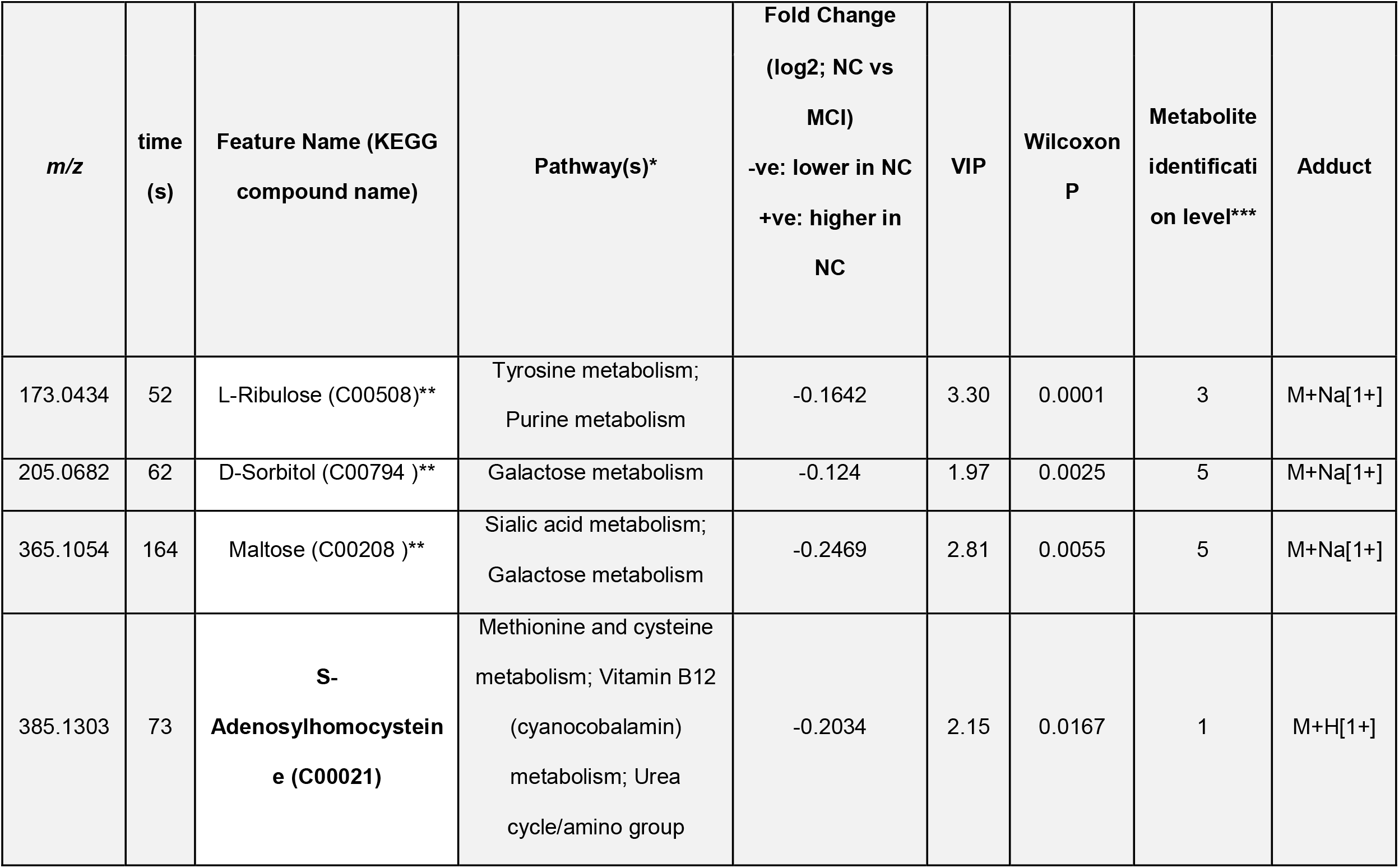
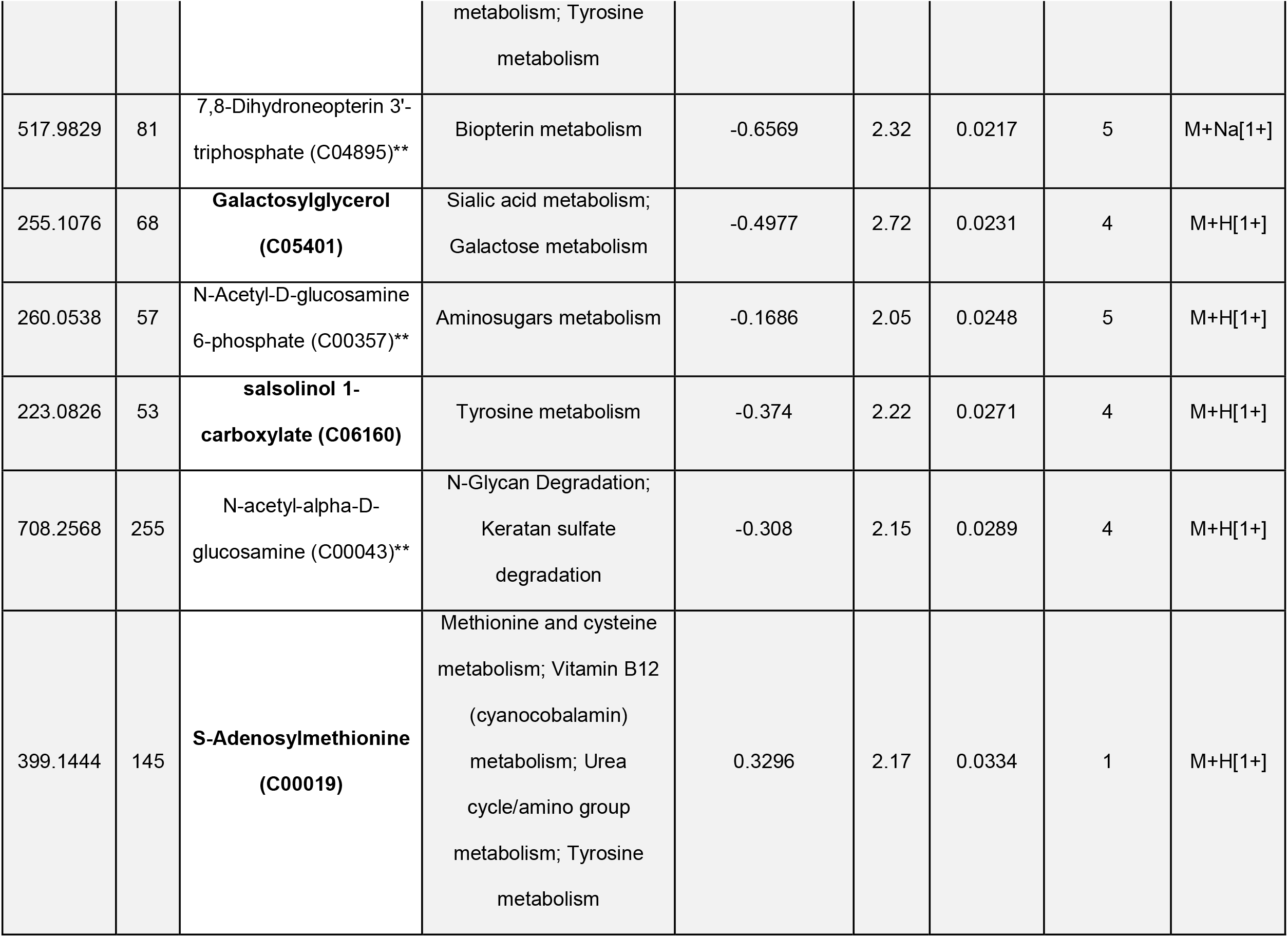
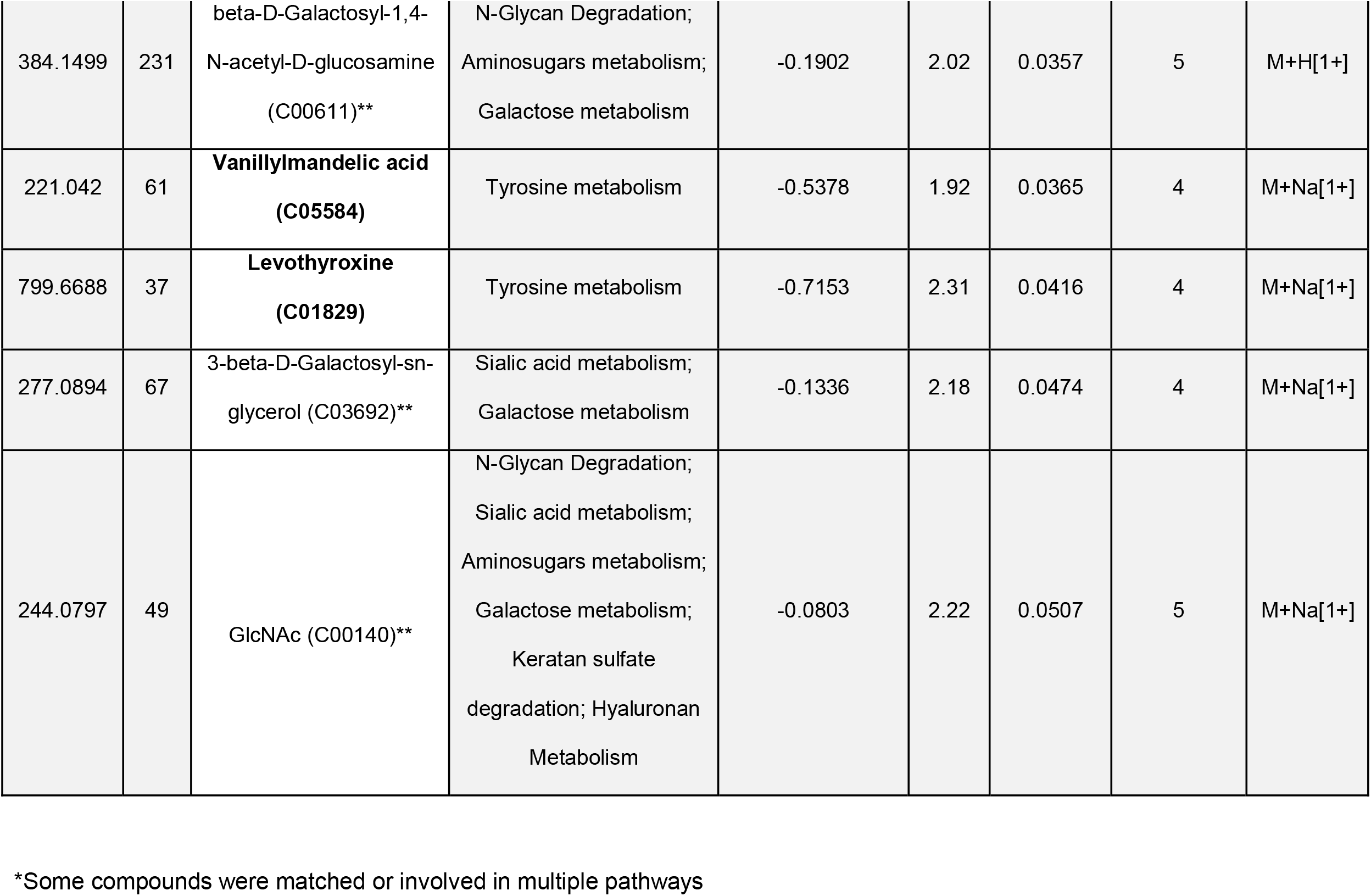
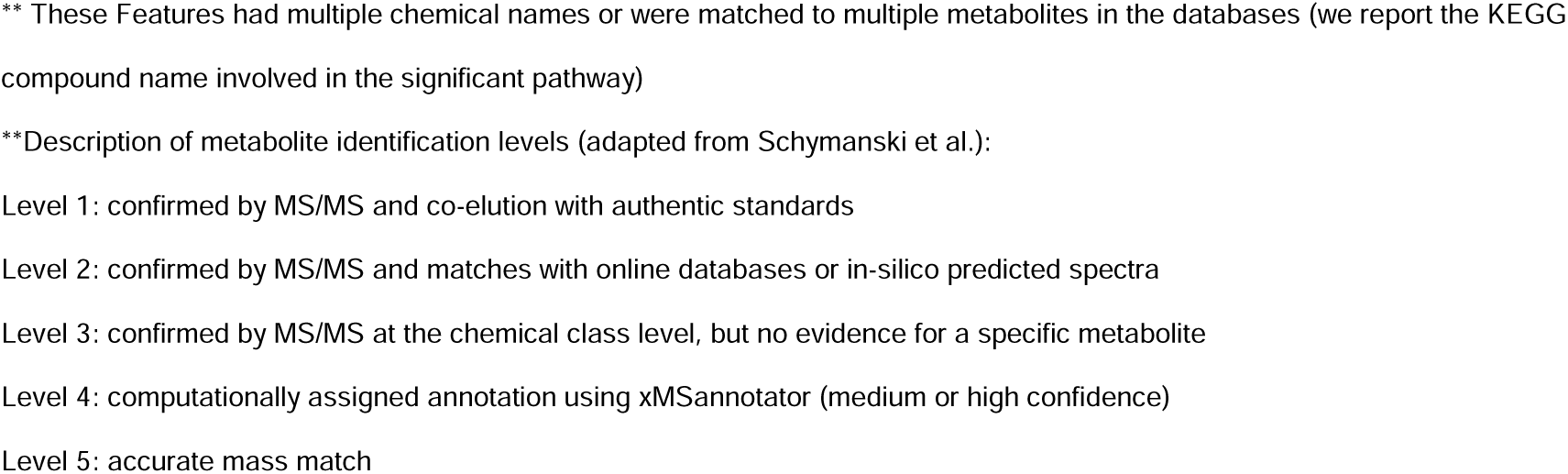
Results of the Pathway Enrichment Analysis with the significant features and associated pathways in the Normal vs MCI groups

| *m/z* | time<br>(s) | Feature Name (KEGG<br>compound name) | Pathway(s)* | Fold Change<br>(log2; NC vs<br>MCI)<br>-ve: lower in NC<br>+ve: higher in<br>NC | VIP | Wilcoxon<br>P | Metabolite<br>identification<br>level*** | Adduct |
| --- | --- | --- | --- | --- | --- | --- | --- | --- |
| 173.0434 | 52 | L-Ribulose (C00508)** | Tyrosine metabolism;<br>Purine metabolism | -0.1642 | 3.30 | 0.0001 | 3 | M+Na[1+] |
| 205.0682 | 62 | D-Sorbitol (C00794)** | Galactose metabolism | -0.124 | 1.97 | 0.0025 | 5 | M+Na[1+] |
| 365.1054 | 164 | Maltose (C00208)** | Sialic acid metabolism;<br>Galactose metabolism | -0.2469 | 2.81 | 0.0055 | 5 | M+Na[1+] |
| 385.1303 | 73 | **S-**<br>**Adenosylhomocysteine (C00021)** | Methionine and cysteine<br>metabolism; Vitamin B12<br>(cyanocobalamin)<br>metabolism; Urea<br>cycle/amino group metabolism; Tyrosine<br>metabolism | -0.2034 | 2.15 | 0.0167 | 1 | M+H[1+] |
| 517.9829 | 81 | 7,8-Dihydroneopterin 3'-<br>triphosphate (C04895)** | Biopterin metabolism | -0.6569 | 2.32 | 0.0217 | 5 | M+Na[1+] |
| 255.1076 | 68 | **Galactosylglycerol**<br>**(C05401)** | Sialic acid metabolism;<br>Galactose metabolism | -0.4977 | 2.72 | 0.0231 | 4 | M+H[1+] |
| 260.0538 | 57 | N-Acetyl-D-glucosamine<br>6-phosphate (C00357)** | Aminosugars metabolism | -0.1686 | 2.05 | 0.0248 | 5 | M+H[1+] |
| 223.0826 | 53 | **salsolinol 1-**<br>**carboxylate (C06160)** | Tyrosine metabolism | -0.374 | 2.22 | 0.0271 | 4 | M+H[1+] |
| 708.2568 | 255 | N-acetyl-alpha-D-glucosamine (C00043)** | N-Glycan Degradation;<br>Keratan sulfate<br>degradation | -0.308 | 2.15 | 0.0289 | 4 | M+H[1+] |
| 399.1444 | 145 | **S-Adenosylmethionine**<br>**(C00019)** | Methionine and cysteine<br>metabolism; Vitamin B12<br>(cyanocobalamin)<br>metabolism; Urea<br>cycle/amino group<br>metabolism; Tyrosine<br>metabolism | 0.3296 | 2.17 | 0.0334 | 1 | M+H[1+] |
| 384.1499 | 231 | beta-D-Galactosyl-1,4-N-acetyl-D-glucosamine (C00611)** | N-Glycan Degradation;<br>Aminosugars metabolism;<br>Galactose metabolism | -0.1902 | 2.02 | 0.0357 | 5 | M+H[1+] |
| 221.042 | 61 | **Vanillylmandelic acid (C05584)** | Tyrosine metabolism | -0.5378 | 1.92 | 0.0365 | 4 | M+Na[1+] |
| 799.6688 | 37 | **Levothyroxine (C01829)** | Tyrosine metabolism | -0.7153 | 2.31 | 0.0416 | 4 | M+Na[1+] |
| 277.0894 | 67 | 3-beta-D-Galactosyl-sn-glycerol (C03692)** | Sialic acid metabolism;<br>Galactose metabolism | -0.1336 | 2.18 | 0.0474 | 4 | M+Na[1+] |
| 244.0797 | 49 | GlcNAc (C00140)** | N-Glycan Degradation;<br>Sialic acid metabolism;<br>Aminosugars metabolism;<br>Galactose metabolism;<br>Keratan sulfate degradation; Hyaluronan Metabolism | -0.0803 | 2.22 | 0.0507 | 5 | M+Na[1+] |

\*Some compounds were matched or involved in multiple pathways
\*\* These Features had multiple chemical names or were matched to multiple metabolites in the databases (we report the KEGG compound name involved in the significant pathway)
\*\*Description of metabolite identification levels (adapted from Schymanski et al.):
Level 1: confirmed by MS/MS and co-elution with authentic standards
Level 2: confirmed by MS/MS and matches with online databases or in-silico predicted spectra
Level 3: confirmed by MS/MS at the chemical class level, but no evidence for a specific metabolite
Level 4: computationally assigned annotation using xMSannotator (medium or high confidence)
Level 5: accurate mass match

**Figure 2:**
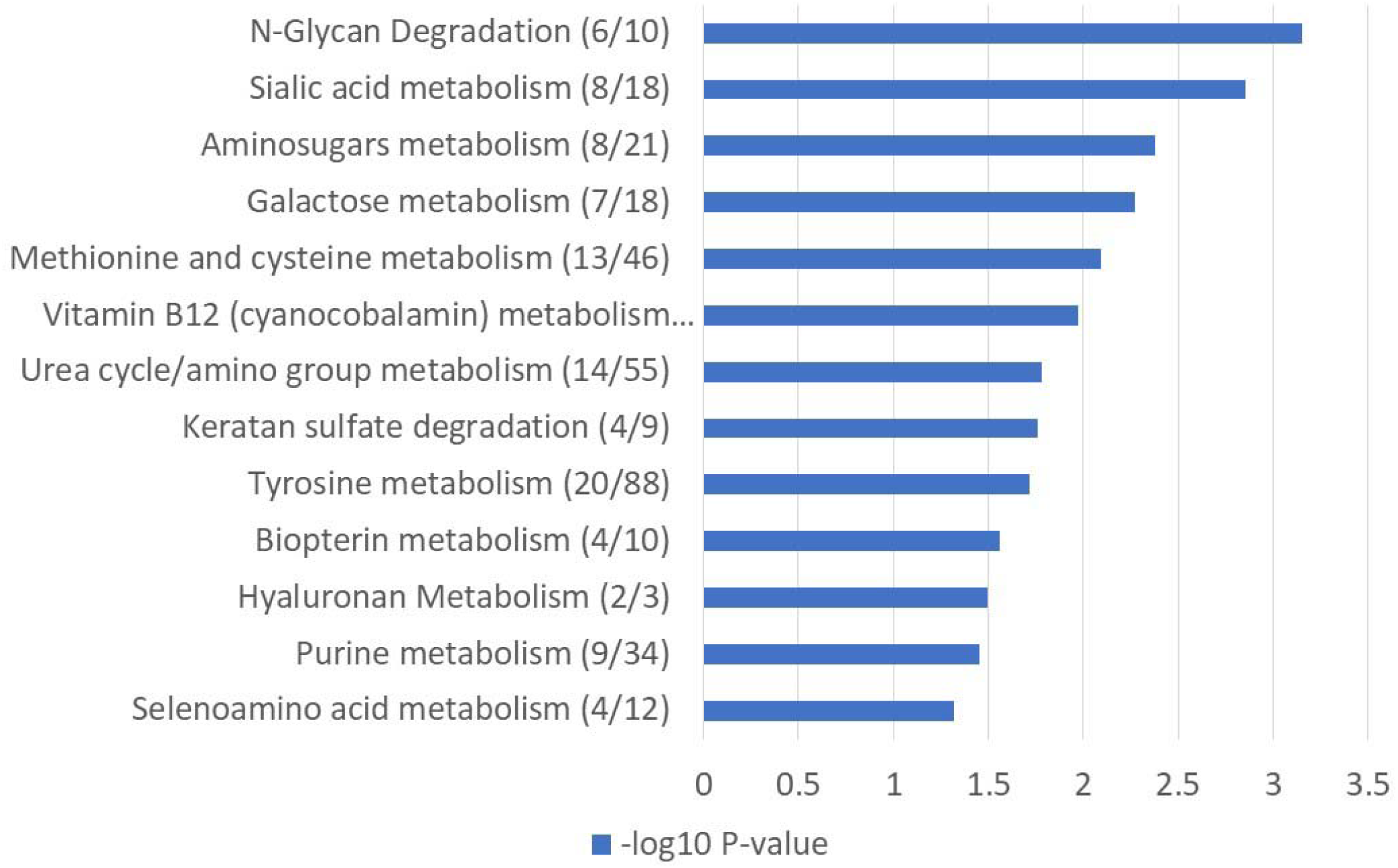
Pathways altered in MCI cases compared to controls. (Pathway analysis was performed using Mummichog 2.0.6 on the 1049 features identified by PLS-DA with a VIP ≥ 1.5.)

**Figure 3:**
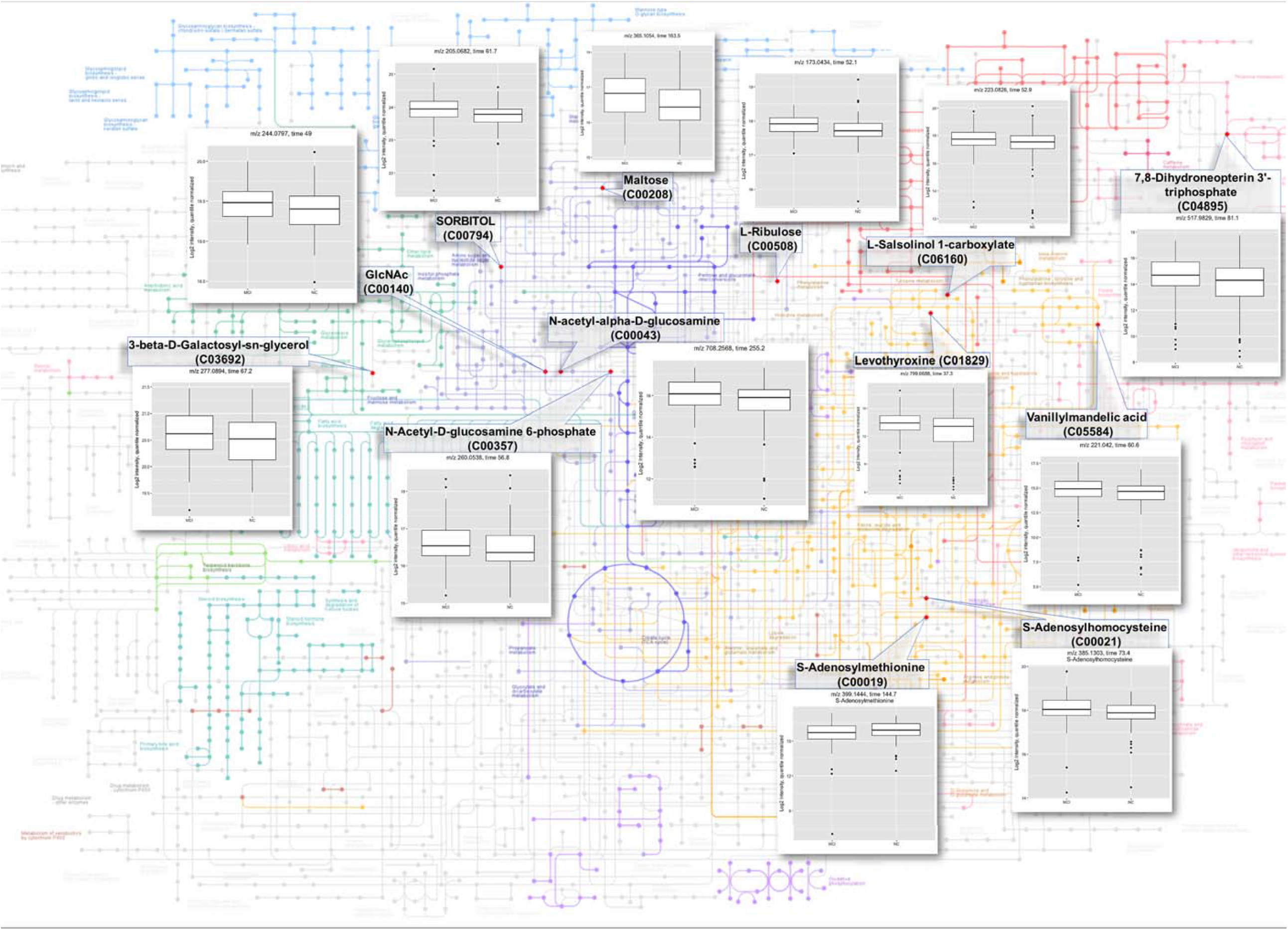
Significant Features identified from the Pathway enrichment analysis and their BOX-PLOTs comparing normal to MCI participants.

### 3.4. Correlation with disease phenotype

We then explored the associations between these signature features with disease phenotypes. These results are shown in **Figure 4**. Increased expression of bioenergetics and glucose metabolism were associated with higher tau and Ptau but also with lower cognitive performance, hippocampal volume and cortical thickness, decreased cognitive and neuroimaging. Sugar metabolism dysregulations were associated with increased Tau, and ptau. Further, 5 of these features were associated with decreased cortical thickness, hippocampal volume and cognitive performance on MoCA, TMT and delayed recall. S-Adenosylhomocysteine was associated with lower MoCA scores and decreased cortical thickness. In the Tyrosine pathway, Salsolinol-1-Carboxylate was associated with higher Tau and pTau whereas VMA was associated with lower MoCA Score. Finally, features in the biopterin pathway were not associated with any disease phenotype. Detailed results are provided in **supplemental Table 3**.

**Figure 4.**
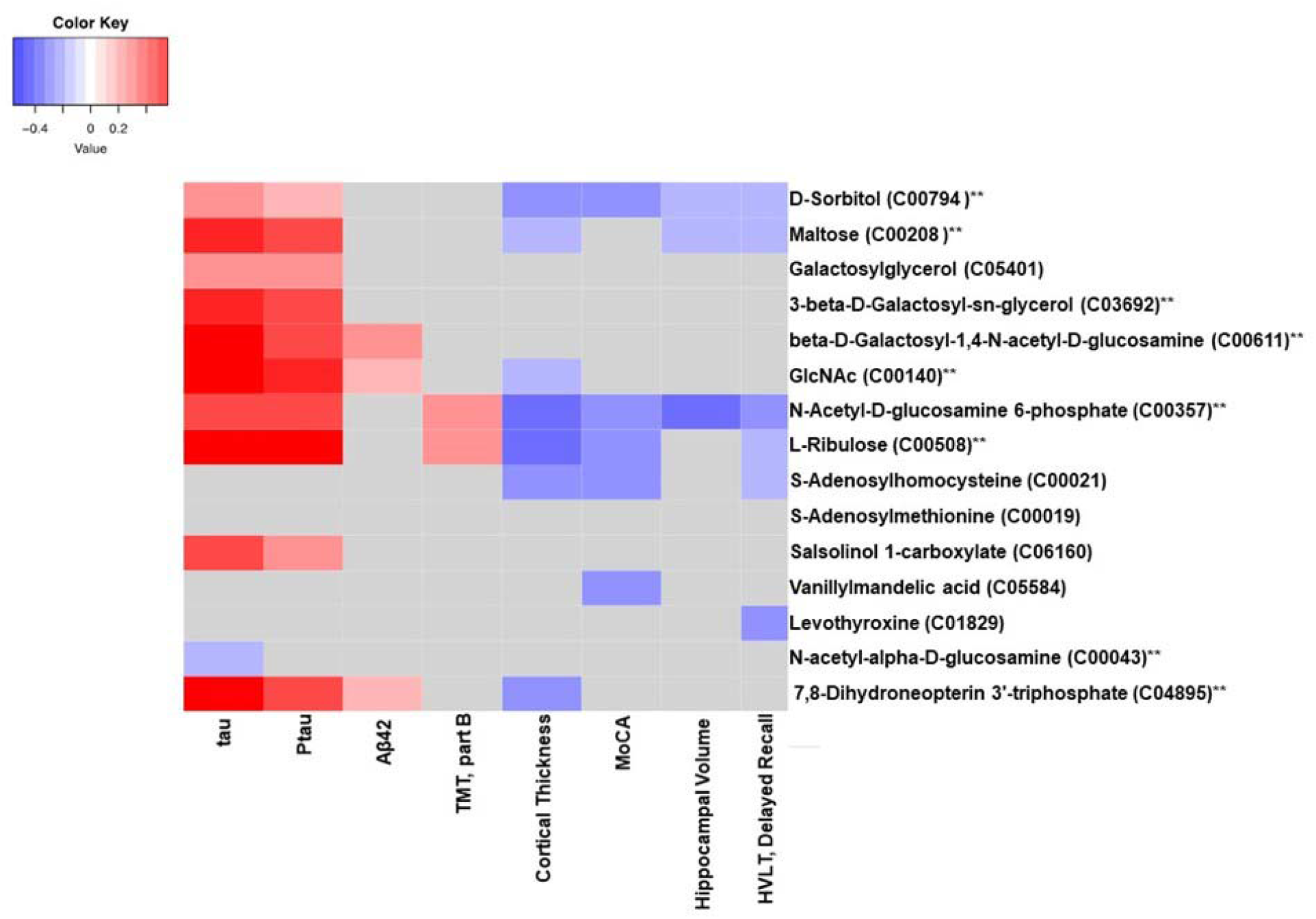
Correlations between significant features and disease phenotypes. Spearman correlation coefficients. Red indicates positive correlations and blue indicates negative correlations. Correlations with *p* ≥0.05 are marked in grey.

## 4. Discussion

This study of untargeted high resolution metabolomics identified a CSF signature in prodromal AD characterized by dysregulation of sugar, homocysteine/methionine and tyrosine metabolism. Multiple features within this signature was associated with increased total tau and Ptau biomarkers and lower cognitive measures, hippocampal volume and cortical thickness.

Multiple studies have suggested an association between AD and peripheral impaired glucose metabolism that may be pronounced in those with type 2 Diabetes and insulin resistance. [37, 38] There is also evidence that in AD, a central insulin resistant state is detectable even in the pre-symptomatic stages.[39, 40] Our study suggests that in the CSF of those with MCI, there was dysregulation of multiple glucose-metabolism pathways and related increase in glucose metabolism byproducts. Prior FDG-PET scans have suggested decreased brain metabolisms across the spectrum of AD.[41, 42] Taken together, the increased in CSF features of sugar metabolism pathways couple with the previously reported brain hypometabolism may in part be explained by a lower brain glucose uptake, for example secondary to glucose uptake transporter pathways, [43, 44] leading to an increased CSF levels. An alternative explanation is that the possible central insulin resistance reported in AD is associated with increases metabolic by-products in the brain and CSF. This is further supported by our observation that these increased metabolic features are associated with increased tau measures and with lower performance on cognitive assessments, hippocampal volume and cortical thickness. The link between the various metabolism and energetic pathways with AD and tau has been reported previously using FDG-pet. [45, 46] We did not measure brain glucose uptake or metabolism and hence these are potential explanations that need further confirmation.

Our observation that alterations in pathways related to homocysteine and methionine metabolisms is of great interest. Specifically, S-adenosylmethionine (SAM) was under-expressed and S-adenosylhomocysteine (SAH) was overexpressed in CSF of the MCI participants. SAM is a key molecule in methionine cycle involved in nucleic acid and protein metabolism and synthesis. SAH is formed by demethylation of SAM. Prior reports suggest that SAM is decreased and SAH is increased in CSF of AD and are related to tau bioamrkers.[47]

However, in this study only SAM was related to additional disease phenotypes including cognitive measures and cortical thickness. Nevertheless, this untargeted approach suggest that homocysteine-methionine pathways are dysregulated in the prodromal stages of AD.

We identified perturbations in Tyrosine pathways with overlapping features in the purine, methionine and homocysteine pathways, including SAM, SAH, VMA, and thyroxine. These cycles are involved in catecholamine and serotonin neurotransmitter systems and might be altered in AD.[48] A prior CSF analysis in a smaller number of MCI using targeted metabolomic approach suggested a similar finding of impairments in methionine and tyrosine pathways.[49] Despite the difference between the groups in this pathway, there were minimal associations with the other disease measures.

There are multiple advantages to this study including the untargeted and advanced bioinformatic approaches which allowed us to consider a large number of pathways and features, the comparably larger number of sample with CSF, and the availability of multiple additional disease phenotypes that offer greater confidence in the associations with MCI. The limitations include the cross-sectional design and the number of identified features that could not be matched to known metabolites or matched to multiple metabolites, which is a major bottleneck in untargeted metabolomics.[6] The use of MS/MS with an in-house library of confirmed metabolites in the Clinical Biomarker Lab where these analyses were performed using authentic standards. This coupled with advanced bioinformatics tools for metabolite identification and annotation enhanced the reliability of the identity of our metabolites compared to many prior untargeted studies.

Clinical translations of these findings are important. The key pathways that are perturbed in AD are potential targets for existing or new drug developments. For example, insulin and other antidiabetic agents may address the sugar metabolism abnormalities identified in this analysis. [50, 51] Drugs that may restore balance between SAM and SAH may also be of relevance in the drug development of AD.[52]

## 5. Conclusion

In this untargeted HRM study, we identified a metabolic signature characterized by impairments in sugar metabolism and methionine, homocysteine and tyrosine pathways in MCI. These offer insight into the metabolic derangements that occur in predementia stages of AD and potential therapeutic targets.

## Data Availability

ALl data will be made available at SYNAPSE in 2020.

https://www.synapse.org/

## Acknowledgement

This research was funded by the National Institute on Aging grants AG051633, AG057470-01, and AG042127 to IH.

This work was supported by the National Institutes of Health [Grants number RF1AG051633, RF1AG057470, R01AG049752, R01AG042127]

Authors have no competing interests to declare

